# *Streptococcus pneumoniae* colonisation associates with impaired adaptive immune responses against SARS-CoV-2

**DOI:** 10.1101/2021.07.22.21260837

**Authors:** Elena Mitsi, Jesús Reiné, Britta C Urban, Carla Solórzano, Elissavet Nikolaou, Angela D. Hyder-Wright, Sherin Pojar, Ashleigh Howard, Lisa Hitchins, Sharon Glynn, Madlen Farrar, Konstantinos Liatsikos, Andrea M Collins, Naomi F Walker, Helen Hill, Esther L German, Katerina S Cheliotis, Rachel L Byrne, Christopher T. Williams, Ana I Cubas-Atienzar, Tom Flecher, Emily R Adams, Simon J Draper, David Pulido, Rohini Beavon, Christian Theilacker, Elizabeth Begier, Luis Jodar, Bradford D Gessner, Daniela M Ferreira

**Affiliations:** Clinical Sciences, Liverpool School of Tropical Medicine, Liverpool, UK; Tropical Disease Biology, Liverpool School of Tropical Medicine, Liverpool, UK; Liverpool University Hospitals NHS Foundation Trust, Liverpool; Jenner Institute, University of Oxford, Oxford, UK; Pfizer Vaccines, Collegeville, Pennsylvania, United States

**Author notes:** Corresponding Authors: Prof Daniela Ferreira and Dr Elena Mitsi, Liverpool School of Tropical Medicine, 1 Daulby Street, Liverpool L7 8XZ, United Kingdom.

**Keywords:** SARS-CoV-2, *S. pneumoniae*, immune responses, healthcare workers, hospital presented patients

## Abstract

Although recent epidemiological data suggest that pneumococci may contribute to the risk of SARS-CoV-2 disease, secondary pneumococcal pneumonia has been reported as infrequent. This apparent contradiction may be explained by interactions of SARS-CoV-2 and pneumococcus in the upper airway, resulting in the escape of SARS-CoV-2 from protective host immune responses. Here, we investigated the relationship of these two respiratory pathogens in two distinct cohorts of a) healthcare workers with asymptomatic or mildly symptomatic SARS-CoV-2 infection identified by systematic screening and b) patients with moderate to severe disease who presented to hospital. We assessed the effect of co-infection on host antibody, cellular and inflammatory responses to the virus. In both cohorts, pneumococcal colonisation was associated with diminished anti-viral immune responses, which affected primarily mucosal IgA levels among individuals with mild or asymptomatic infection and cellular memory responses in infected patients. Our findings suggest that *S. pneumoniae* modulates host immunity to SARS-CoV-2 and raises the question if pneumococcal carriage also enables immune escape of other respiratory viruses through a similar mechanism and facilitates reinfection occurrence.

## INTRODUCTION

Despite the widespread global effects of the coronavirus disease 2019 (COVID-19) pandemic, few reports have assessed potential interactions between upper airway bacterial colonisation and the severe acute respiratory syndrome coronavirus 2 (SARS-CoV-2). Consequently, the contribution of respiratory bacterial pathogens to SARS-CoV-2 infection and pathogenesis remains poorly understood^1, 2^. However, post-hoc analysis of two randomised clinical trials has found that individuals vaccinated with pneumococcal conjugate vaccines (PCVs) showed a reduction of 30-35% in hospitalisations for endemic human coronavirus (HCoV, OC43 and HKU1) associated pneumonia in adults^3, 4^ and lower respiratory infection in children^5^. A recent observational study reported that 13-valent pneumococcal conjugate vaccine (PCV13) in older adults was associated with a reduction of approximately 30% in COVID-19 disease, hospitalisation, and death^6^. Also, recent epidemiological studies report higher mortality observed in patients with SARS-CoV-2 co-infection or subsequent infection (although rare events) within 28 days after invasive pneumococcal disease (IPD) in the UK^7^. Traditionally, viral-pneumococcal interaction in the upper airway has been thought to increase the risk of secondary pneumococcal pneumonia, particularly during influenza and RSV seasonal outbreaks^8^. However, a low proportion of COVID-19 patients have had documented pneumococcal pneumonia based on blood and sputum culture^1, 7, 9^. Reasons for lower occurrence of secondary pneumococcal pneumonia compared to previous influenza pandemics may be low pneumococcal transmission rates due to non-pharmacologic interventions, such as reduced social mixing, or low detection rates by microbiologic culture due to liberal empiric antibiotic use in the severely ill patient population^1, 7^.

Bacterial and viral interaction in the upper airways could confer protection against viral infection by priming the immune response or act synergistically to promote viral evasion by direct and indirect mechanisms^10, 11^. Adaptive immune mechanisms play a critical role in protecting against viral infection, including against SARS-CoV-2^12–14^. *S. pneumoniae* has been shown to have a modulatory effect on the anti-viral immune responses mounted by the host and the sequence of pathogen exposure in co-infection cases may also alter the disease outcome. Mice exposed to *S. pneumoniae* prior to influenza A exhibited reduced antiviral serum IgG with diminished virus neutralisation activity a month after infection^15^. A randomised controlled human study of experimental pneumococcus/influenza co-infection reported diminished IgA responses to influenza antigens associated with pneumococcal carriage^16^, resembling findings of the current study. Non-pneumococcal specific cleavage of mucosal IgA1 by pneumococcal IgA1 proteases^17^ could be a potential mechanism which contributes to anti-viral IgA reductions identified in both studies.

To study interactions of pneumococcus and SARS-CoV-2 and the effect of pneumococcus on host anti-viral immune responses, we longitudinally sampled a cohort of healthcare workers (HCW) at high risk for SARS-CoV-2 infection and patients with suspected COVID-19 disease. In both cohorts, we studied prevalence of SARS-CoV-2 and pneumococcal colonisation, associations of co-infection and disease severity and evaluated immune responses and inflammation levels in the context of SARS-CoV-2 mono-infection and co-infection with pneumococcus. Lastly, we sought to assess whether pneumococcal carriage associated reduction in mucosal IgA to respiratory viruses could be due the activity of pneumococcus IgA1 protease as well as whether the order of infection of virus and pneumococcus was important. Consequently, we evaluated samples from our previous studies of live attenuated influenza virus vaccine (LAIV) and pneumococcus co-infections ^11, 16, 18^ to assess that.

## RESULTS

### SARS-CoV-2 and *S. pneumoniae* prevalence in HCW and Patient Cohort

The impact of pneumococcal carriage status on SARS-CoV-2 viral replication and clinical outcome was assessed in a cohort of frontline HCWs (n=85, median age: 35; IQR: 27.5-46.5) and a cohort of patients presented to hospital with suspected COVID-19 disease (patients, n=400, median age: 61; IQR:48 -72). Participants from these prospective studies were screened for both SARS-CoV-2 and *Streptococcus pneumoniae (Spn)* presence in the naso/oropharynx (Figure 1A). Amongst HCWs, 34% (29/85) tested positive for SARS-CoV-2 at any time point during the 3-months follow-up period of the study on a nasopharyngeal (NP) swab or NP swab and saliva sample and all of these experienced asymptomatic or mildly symptomatic viral infection^19^. In the patient cohort, 63.5% (255/400) were tested positive for SARS-CoV-2 at the point of recruitment to the study on NP swabs, and their symptoms ranged from moderate to severe^20^. In the HCW cohort, the overall Spn colonisation rate was 20% (17/85). Increased Spn prevalence was observed in SARS-CoV-2 positive compared to the SARS-CoV-2 negative participants [34.5% (10/29) vs 12.5% (7/56), respectively, p=0.023] (Figure 1B), with 7/10 participants acquiring SARS-CoV-2 while already being colonised with Spn and 3/10 having a concurrent infection. In the patient cohort, the overall Spn colonisation rate was 8.5% (35/400) and prevalence of Spn colonisation did not differ amongst SARS-CoV-2 positive and SARS-CoV-2 negative individuals [9.4% (24/255) vs 7.6% (11/145), respectively] (Figure 1C). Spn colonised and non-colonised patients did not differ in disease severity, as defined by NIH severity score (median: 4, IQR:3-4 in both groups), or in days of sample collection post symptoms onset (median: 41, IQR:29-57 vs 47, IQR: 36-59) (Table 1). However, recruitment of patients who had already developed symptoms made the order of pathogen infection unknown. During the 9 months recruitment period of patient study, we observed fluctuations in pneumococcal carriage rate, with lower colonisation rates during periods of local and UK lockdowns (Figure 1D).

**Figure 1.**
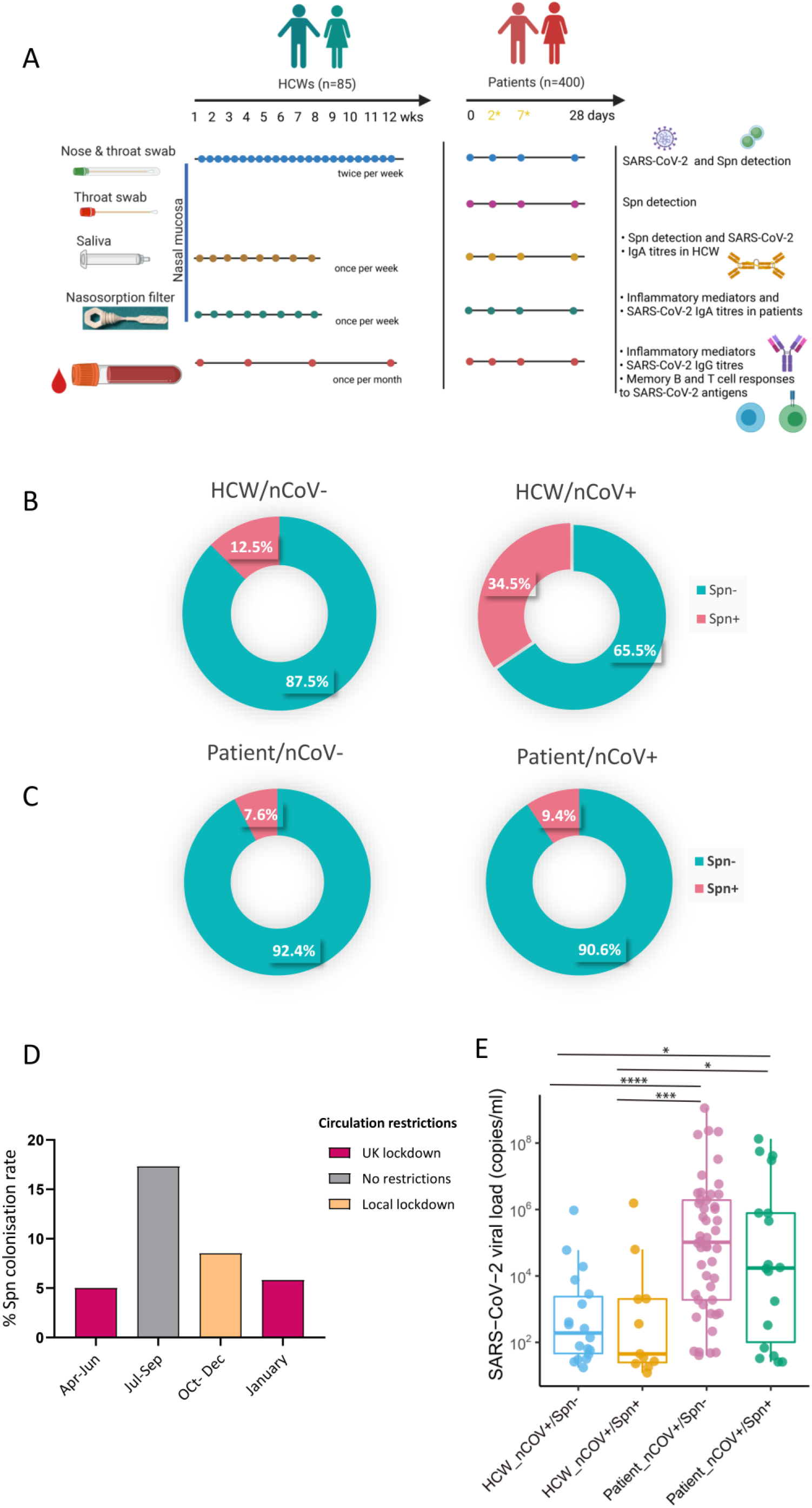
Prevalence of pneumococcal colonisation amongst SARS-CoV-2 positive and negative HCWs and patients. **A)** Experimental design of the study with sample type, sample collection schedule and measurable per sample type depicted for both HCW and patient cohorts. In patient cohort, day2 and day7 samples were collected only if individuals who were hospitalised. **B-C**) Doughnut charts showing the percentage of pneumococcal prevalence in **B)** HCWs (n=85) and **C)** patients (n=400), infected and non-infected with SARS-CoV-2. Fisher’s exact test was used to compare percentages. **D)** Percentage of pneumococcal colonisation rate detected in patient cohort during calendar periods of different circulation restrictions rules applied. 5% (6/119) from April-June, 17.3% (13/75) from July to September, 8.5% (13/154) from October to December and 5.8% (3/52) in January. **E)** Levels of viral load, expressed as RNA copies/ml as detected by Genesig RT-qPCR in HCW non-colonised (n=19, light blue) and Spn-colonised (n=10, yellow) and patient non-colonised (n=73, lilac) and Spn-colonised (n=19, green).

**Table 1:**
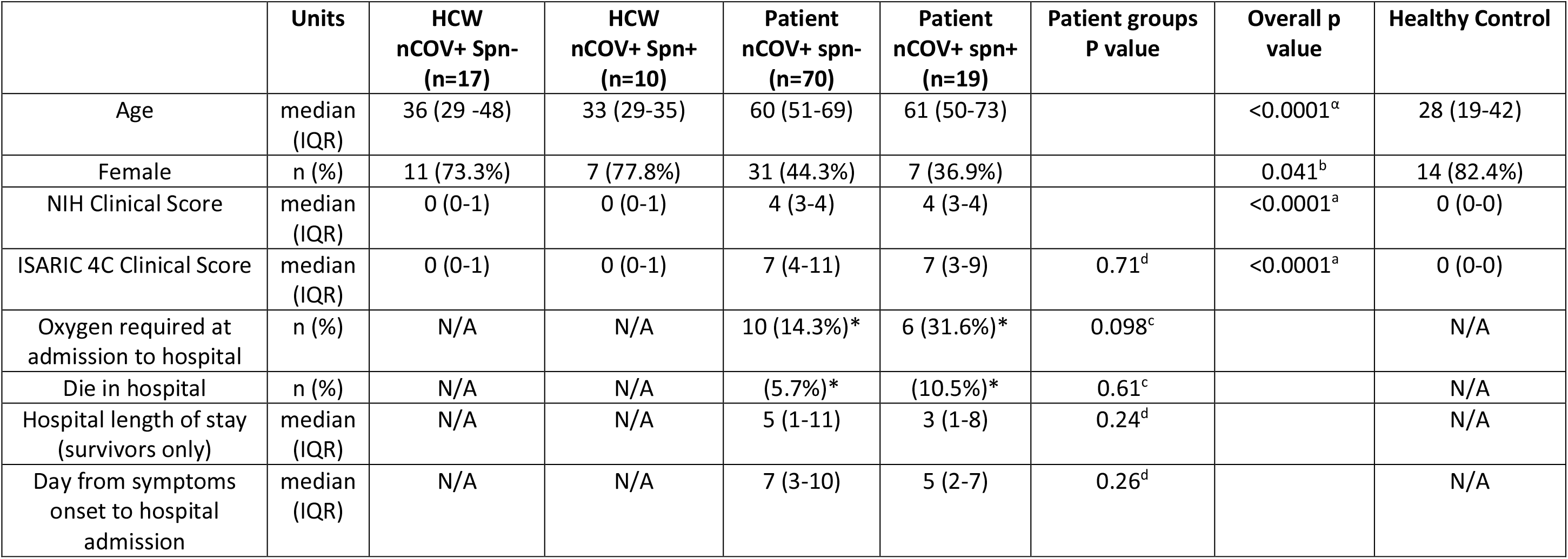
Demographic and clinical Characteristics of the study groups, used in the immunological analysis. Health care workers with RT-qPCR confirmed COVID-19 infection, colonised or non-colonised with pneumococcus. Patients presented to the hospital with RT-qPCR confirmed COVID-19 infection, colonised or non-colonised with pneumococcus. Healthy controls were samples collected from human studies before 2019. * Positivity proportion was calculated using the denominator for individual variables. NIH; National Institutes of Health clinical score for assessment of clinical spectrum of SARS-CoV-2 infection. a: Kruskal-Wallis test, b: Chi-square test, c: Fisher’s exact test, d: Mann-Whitney test.

The SARS-CoV-2 upper airway viral load did not differ significantly by Spn carriage status in either the HCW (median: 2.01 x10^2^ RNA copies/ml, IQR: 4.02 x10^1^-4.03 x10^3^ in Spn-vs 4.5 x 10^1^ RNA copies/ml, IQR: 2.30 x10^1^-2.03 x10^3^ in Spn+) or the patient cohort (median: 1.04 x 10^5^ RNA copies.ml, IQR: 1.89 x10^3^-1.94 x10^6^ in Spn- vs 1.74 x 10^4^ RNA copies/ml, IQR: 6.12 x10^1^- 8.14 x10^6^ in Spn+). The patient cohort (both Spn colonised and non-colonised groups) had a higher viral load compared to the HCW cohort (Figure 1E).

### Impaired mucosal antibody responses to SARS-CoV-2 in pneumococcal colonised individuals

IgA plays a crucial role in the immune defence of mucosal surfaces, the first point of entry of SARS-CoV-2^13, 21, 22^. We measured levels of mucosal IgA to surface SARS-CoV-2 antigens (Receptor Binding Domain; [RBD], spike protein subunit 1; [S1], and spike protein subunit 2; [S2]) but also to the internal nucleocapsid protein (N) in saliva samples in HCWs and nasal lining fluid in patient cohort (due to difficulties in acquiring saliva from patients) one month post SARS-CoV-2 infection.

In the HCW cohort, among SARS-CoV-2 positive subjects, Spn colonised HCW had lower salivary IgA levels than non-colonised HCWs, with statistically significant differences for S1 and S2 between the two HCW groups for all SARS-CoV-2 antigens assessed (median 4.1- and 6.4-fold change of IgA to S1 and S2 from control levels, respectively) (p=0.012 and p=0.009, respectively) (Figure 2A). Non-colonised participants but not the Spn colonised group mounted robust salivary IgA responses to all four viral antigens. Spn colonised HCWs elicited IgA against RBD and N but not to S1 and S2 antigens, which levels were similar to uninfected healthy controls (Figure 2A).

**Figure 2.**
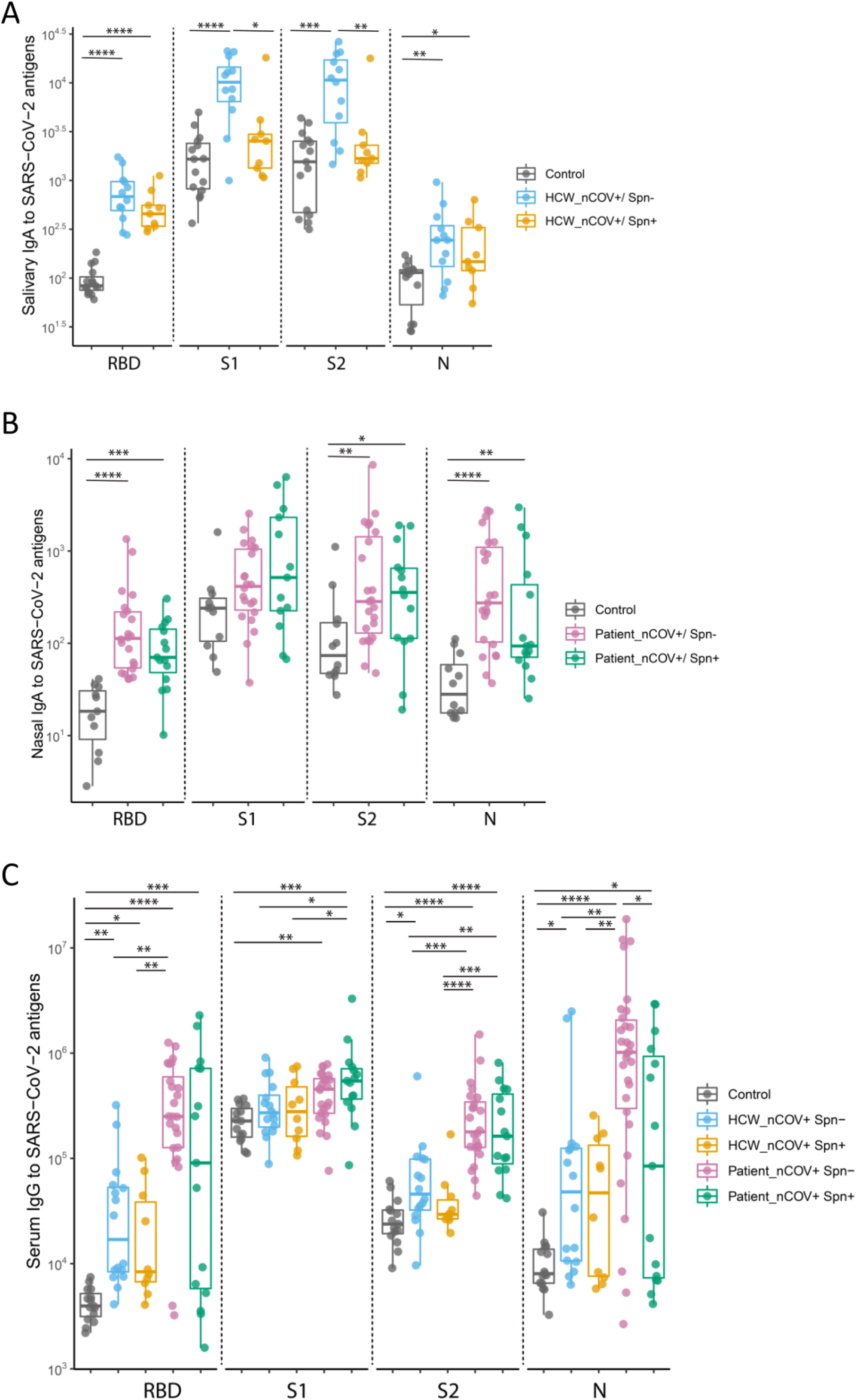
Mucosal and systemic antibody responses to SARS-CoV-2 in HCWs and patients. **A)** Salivary IgA titres to SARS-CoV-2 RBD, S1, S2 and N protein in HCWs, divided into non-colonised (n=12) and Spn-colonised (n=9), and unexposed healthy controls (n=15). **B)** Nasal IgA titres to SARS-CoV-2 RBD, S1, S2 and N protein in patients, divided into non-colonised (n=23) and Spn-colonised (n=15) and unexposed healthy control collected before 2019 (n=12). **C)** Serum IgG titres in HCW (non-colonised; n=16 and Spn-colonised; n=10), patients (non-colonised, n=24 and Spn-colonised, n=14) and unexposed healthy controls (n=15). Both mucosal and serum antibody titres from SARS-CoV-2 positive participants were measured during the convalescent phase of the viral infection. Antibody levels are expressed as arbitrary units. Medians with IQRs are depicted for anti-viral responses. *p <0.05, **p<0.01, ***p < 0.001, ****p < 0.0001 by Mann-Whitney test (panels A and B) or by Kruskal-Wallis test (panel C) for comparisons between groups.

In the patient cohort, among SARS-CoV-2 positive subjects, despite a trend of overall weekend IgA induction to SARS-CoV-2 antigens in Spn colonised patients compared to non-colonised patients, there were no significant differences between the groups (Figure 2B). Both Spn colonised and non-colonised patients mounted robust nasal IgA to SARS-CoV-2 antigens (RBD, S2 and N), with the exception of S1, of which titres did not differ significantly from control in either group (p=0.053; non-colonised and p=0.093; Spn colonised) (Figure 2B). Particularly, non-colonised patients had the highest antibody fold rise against RBD and N (6.2-fold and 9.4-fold increase from control levels) (p<0.0001 in both), whereas the pneumococcal colonised patient group showed a lesser increase against them (2.2- and 3.2-fold increase from control levels) (p=0.0002 and p=0.0017, respectively) (Figure 2B).

Anti-viral IgG responses were measured in convalescent sera in both cohorts. In the HCW cohort, despite a trend of overall lower levels of IgG induction observed for RBD and S2 antigens in the Spn colonised participants compared to non-colonised participants, there were no significant differences between the groups (Figure 2C). Non-colonised participants showed a moderate rise in IgG titres against all SARS-CoV-2 proteins, except S1, whereas the Spn colonised counterparts mounted lower IgG responses against those viral antigens, with only anti-RBD IgG levels differing significantly from those observed in healthy controls. (Figure 2C). In the patient cohort, IgG levels against N protein were significantly greater in the non-colonised patients when compared with those mounted by the Spn colonised counterparts (median 12-fold difference, p=0.014) and 2-fold higher against RBD in non-colonised vs Spn colonised group (p=0.10) (Figure 2C). Non-colonised and colonised individuals raised similar IgG levels to the spike subunits but had differential IgG responses to RBD and N proteins. In agreement with findings that disease severity correlates with increased levels of systemic IgG to SARS-CoV-2^23^, we observed that IgG titres to viral antigens were consistently higher in patients than HCWs (Figure 2C).

### Experimentally induced pneumococcal colonisation impairs nasal IgA against influenza antigens but only when colonisation precedes viral infection

We have previously observed similar findings of reduced IgA responses to a respiratory viral infection in Spn colonised subjects. For that study, mucosal IgA responses to influenza antigens but not IgG and IgM was reduced when subjects were experimentally colonised with pneumococcal serotype 6B three days before administration of live attenuated influenza vaccine (LAIV)^16^. Those findings in addition to the ones presented here implied the potential involvement of the pneumococcus IgA1 protease-a cell-associated enzyme which cleaves human IgA1, but not IgA2 ^17^- in the reduction of anti-viral mucosal IgA. To test this hypothesis, we sought to assess the association of pneumococcal carriage with virus specific IgA1 vs IgA2 levels in nasal mucosa after pneumococcal/viral co-infection. We also wanted to assess if the order of virus/pneumococcus infection was important for the reduced antibody induction observed in Spn colonised subjects. For this we used samples previously collected in two LAIV-Spn co-infection studies as the timing for virus and Spn infection were well characterised: LAIV was administered either 3 days after (cohort 1) or 3 days after (cohort 2) Spn ^11, 18^.

Influenza-specific IgA, IgA1 and IgA2 levels were measured in nasal wash samples at baseline and D24 after LAIV administration for a subset of participants (15 Spn+ and 15 Spn-) that received LAIV 3 days after pneumococcus. While no to little induction of total IgA and IgA subclasses to influenza antigens were observed from baseline levels in Spn colonised participants, non-colonised group exhibited a median 3-, 1.9- and 1.7-fold rise of influenza specific IgA, IgA1 and IgA2 titres, respectively (Figure S1A). When the order of infection was inverted (LAIV infection occurred 3 days before pneumococcal challenge)^18^ levels of influenza-specific IgA did not differ between Spn colonised and non-colonised participants (IgA median 1.5- and 1.6-fold increase, respectively) (p=0.28) (Figure S1B). Due to insufficient nasal material from the current study cohorts we could not assess IgA subclass in those samples.

### *S. pneumoniae* colonisation is associated with decreased levels of memory B cells to SARS-CoV-2

Memory B cells are of great importance for long term humoral immunity. To identify SARS-CoV-2 specific memory B cells, fluorescently labelled S1 and S2 antigens were used in peripheral blood mononuclear cells (PBMCs) from HCWs and recovered patients. Antigen-binding memory B cells were defined as CD27^+^ and/or IgD^-^ (Figure S2). Overall, Spn colonised participants showed a trend of reduced frequency of memory B cells to SARS-CoV-2 antigens compared to their non-colonised counterparts, which was observed in both HCW and patient cohorts (Figure 3). In HCW cohort, the proportion of S1-specific memory B cells was significantly higher from healthy controls in non-colonised (0.12% vs 0.01%, p=0.003) and Spn colonised group (and 0.06% vs 0.01%, p=0.031) but less pronounced in the latter group (Figure 3A). In the patient group, non-colonised individuals had the highest frequencies of S1- and S2-specific memory B cells when compared to Spn colonised participants and any other group. More specifically, non-colonised patients had significantly greater proportion of S1-specific memory B cells (0.18% vs 0.08%, p=0.027) and a strong trend of higher S2-specific memory B cell proportion compared to Spn-colonised patients (0.35% vs 0.14%, p=0.09) (Figure 3B).

**Figure 3.**
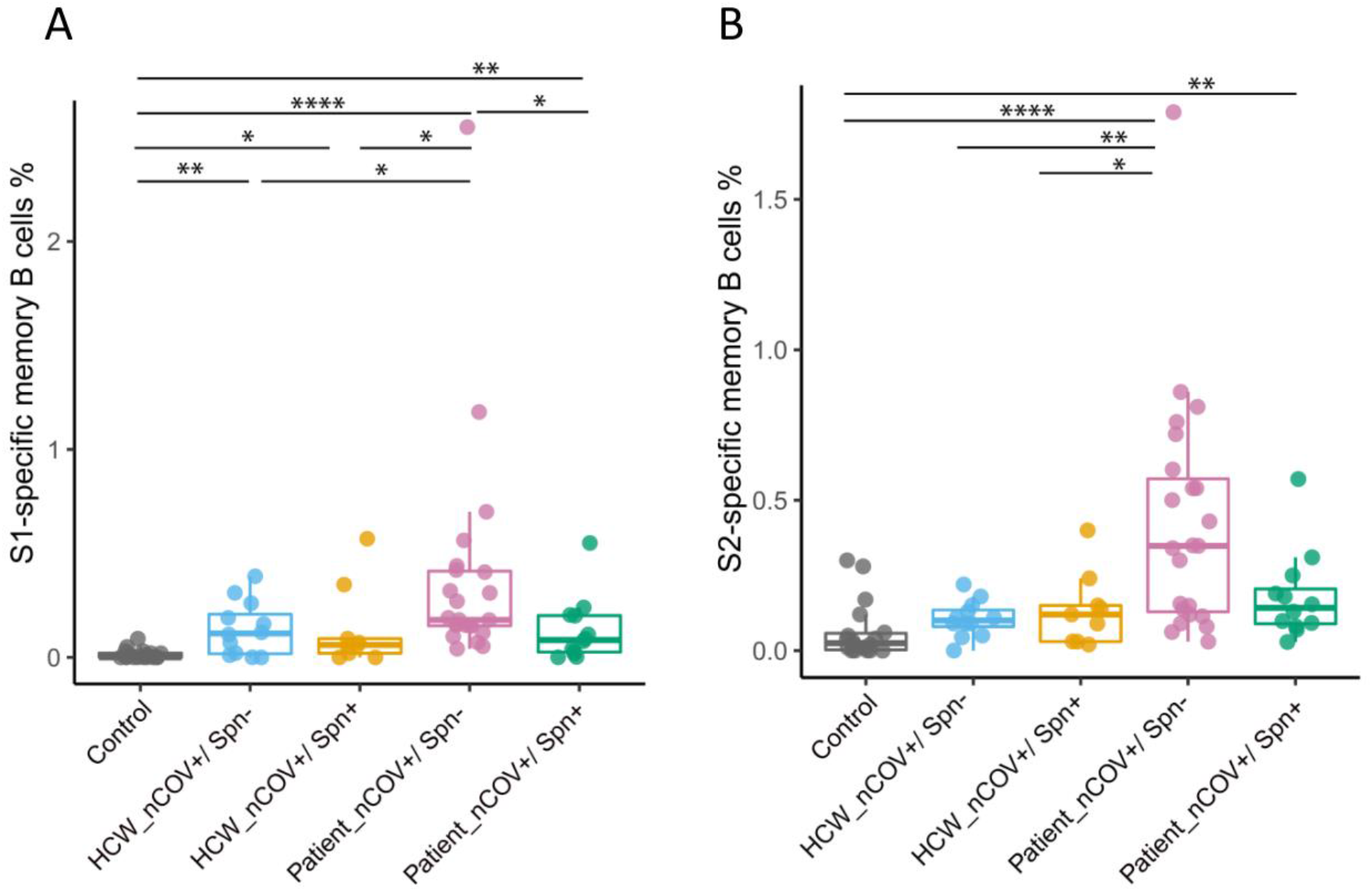
SARS-CoV-2 specific memory B cells in HCW and patients. Percentage of **A)** S1 and **B)** S2 specific memory B cells, within CD19+CD27+ memory B cells in HCW (non-colonised; n=12 and Spn-colonised group; n=9), recovered patients (non-colonised; n=23 and Spn-colonised; n=12) and healthy controls (n=18). Medians with IQRs are depicted and each spot represents an individual. *p <0.05, **p<0.01, ***p < 0.001, ****p < 0.0001 by Kruskal-Wallis test.

### Lack of SARS-CoV-2 specific T cell responses in patients colonised with *S. pneumoniae*

To assess CD4^+^ and CD8^+^ T cell (Figure S3) mediated recall responses in HCWs and convalescent patients, as well as healthy uninfected controls, PBMCs were stimulated *ex vivo* with nucleocapsid (N), spike (S) and subunit 1 (S1) defined peptide pools from SARS-CoV-2. In the HCW cohort, overall CD4^+^ T cells responses did not differ significantly from healthy controls (Figure 4A), either between Spn colonised and non-colonised HCWs. In patient cohort, the magnitudes of T cell responses to N, S1 and S were greater in the non-colonised compared to Spn colonised group and one of the highest in both study cohorts. Median percentages of specific CD4^+^ T cells for N, S1 and S were 1.64% (IQR: 0.53- 2.77), 0.22 (IQR: 0.08-0.54) and 0.57 (IQR: 0.41-1.09), respectively in the non-colonised patient vs 0.14% (IQR: 0.07-0.22), 0.08% (IQR: 0.018-0.38) and 0.12% (IQR: 0.02-0.60), respectively in the Spn-colonised patients. IL-2 was the most abundantly produced cytokine, and similar secretion patterns were observed for TNF-α and IFN-γ, as described above. The cytokine specific (IFN-γ, ΤNF-α and IL-2) CD4^+^ T cells responses to each peptide per group are shown in Figure S4.

**Figure 4.**
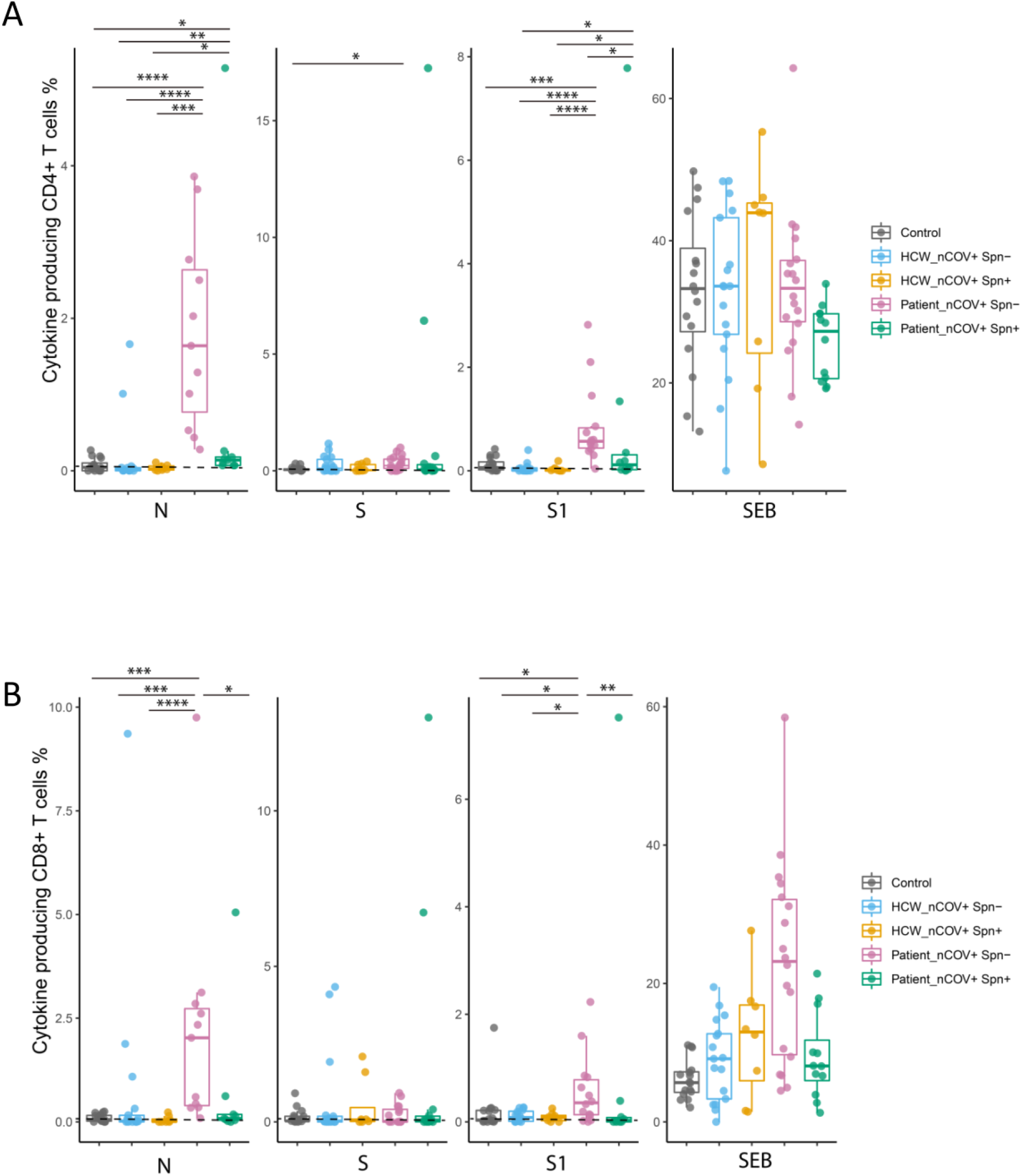
SARS-CoV-2 specific T cell responses in HCW and patients. Percentage of **A)** cytokine producing (IFN-γ, TNF-α, IL-2) CD4+ and **B)** CD8^+^ T cells after *ex vivo* PBMC stimulation with N, S1 and S peptides pools in SARS-CoV-2 positive HCWs (non-colonised; n=17 and Spn-colonised; n=8), recovered patients (non-colonised; n=17 and Spn-colonised; n=14) and healthy control (n=16). One peptide pool was used per condition. SEB was used as a positive control and DMSO as the negative control (non-stimulated cell condition-mock). Background (mock) was subtracted from peptide-stimulated conditions to remove non-specific signal. Medians with IQRs are depicted and each spot represents an individual. *p <0.05, **p<0.01, ***p < 0.001, ****p < 0.0001 by Kruskal-Wallis test.

SARS-CoV-2 N, S and S1 specific CD8^+^ T cell responses were also assessed in the same samples. Similar to CD4^+^ T cell responses, both the number of responders and magnitudes of CD8^+^ responses to N and S1 were the highest and most robust, respectively, in the non-colonised patients. In this group, median CD8^+^ T cell responses for N and S1 were 2.03% (IQR: 0.35%-2.85%) and 0.36% (IQR: 0.11%-0.84%), respectively (Figure 4B). CD8^+^ T cell responses to S peptide pool were close to the lower limit of detection (LOD) in all groups. CD8^+^ responses to N and S1 were impaired in the Spn colonised patients and differed significantly from their non-colonised counterparts (median: 0.08% vs 2.03%, p=0.019 and median: 0.025% vs 0.36%, p=0.009, respectively) (Figure 4B).

### Distinct nasal inflammation profile between HCWs and patients with COVID-19 infection

SARS-CoV-2 induced nasal and systemic inflammatory responses were assessed by measuring levels of 30 cytokines in the nasal fluid and serum during the early phase of the viral infection (acute phase) in HCW and patient cohorts. Nasal and serum samples collected from uninfected adults were used as healthy controls. In blood, non-colonised patients exhibited an increased inflammatory profile (15/30 cytokines were upregulated) compared to the Spn colonised counterparts (upregulation of 9/30 cytokines) and HCWs. In both compartments, HCW groups had a milder pro-inflammatory response compared to patient groups (Figure 5A). Interestingly, in the nose, HCW groups showed a lack of upregulation in cytokines which functionally promote T and B cell maturation and differentiation (Figure 5A). It is important to note, IL-2 and IL-12, which are key cytokines for T cell proliferation and activation, did not differ from control in both HCW and patient Spn colonised groups (Figure 5A). Levels of nasal and serum cytokines per group were also plotted based on significance and fold-change difference from the control group (Figure 5B).

**Figure 5.**
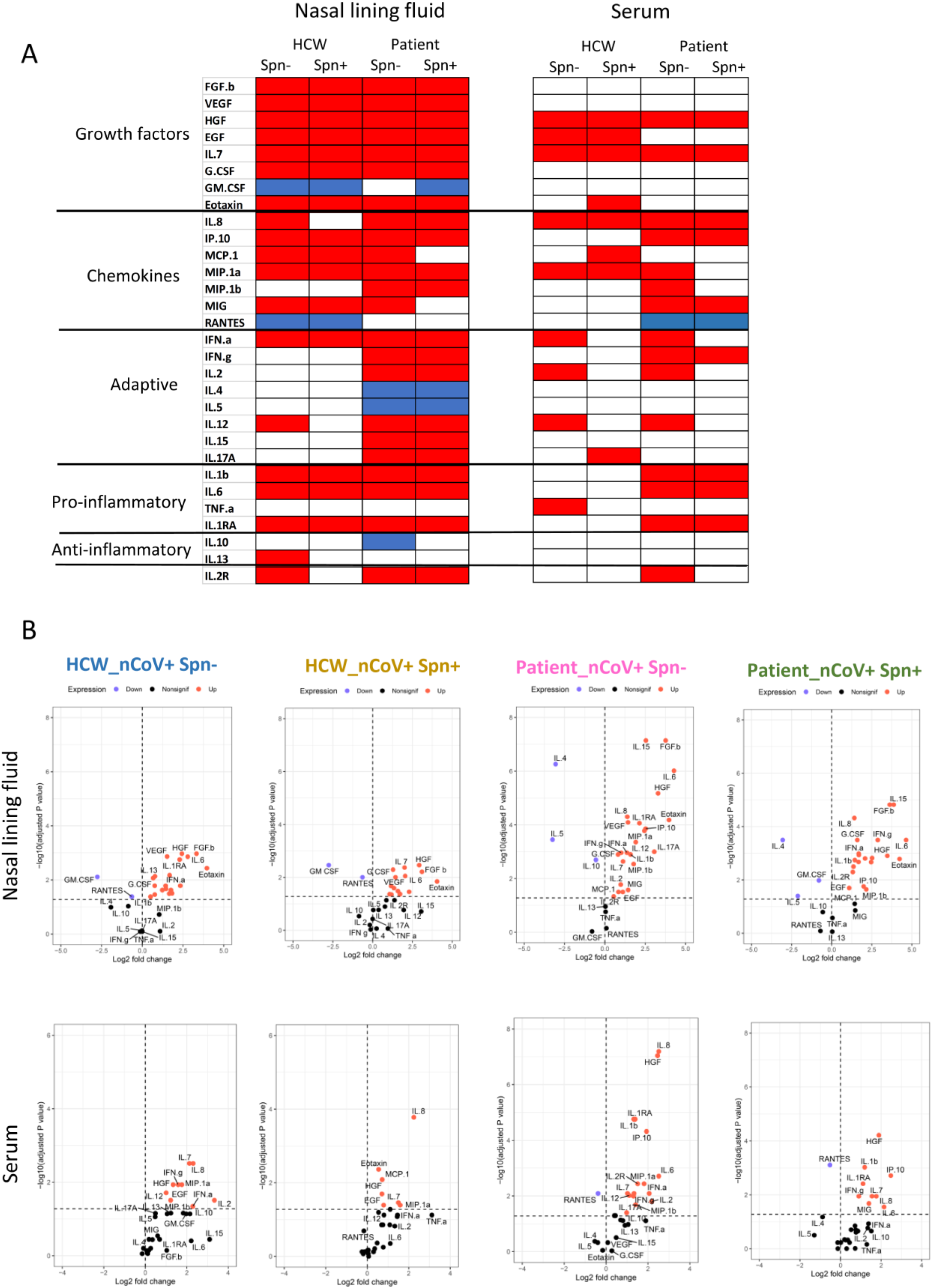
Cytokine concentrations in nasal lining fluid and serum. A) Heatmaps showing increase (red), decrease (blue) or no-significant change of 30-cytokine levels from unexposed healthy control group in nasal lining fluid and serum of non-colonised and Spn-colonised HCWs and patients during the acute phase of SARS-CoV-2 infection. Cytokines were clustered in active cytokine families. B) Volcano plots showing median log2 fold-change from healthy control (n=17) per cytokine in nasal lining fluid and serum of non-colonised HCW(n=17) and Spn-colonised HCW (n=9), non-colonised patient (n=70) and Spn-colonised patient group (n=19). The horizontal dotted line represents the cut-off of significance (p adjusted=0.05, after correction of p value for false discovery rate), while the vertical dotted line represents a cut-off point for determining whether the levels of cytokines were higher (right, red) or lower (left, blue) compared to healthy control group. Statistical comparisons were applied between each study group and the healthy control group using Mann-Whitney test, following Benjamini-Hochberg correction for multiple testing.

We also used an unsupervised analysis to assign profiles to each group. Principal-component analysis (PCA) was applied on all analytes in nasal lining fluid (Figure 6A) and serum (Figure 6B) for all groups and the control. In the nose, patient groups were segregated together and away from the healthy control group in the second principal component (Figure 6A), with the non-colonised group having a more distinctive profile. HCW groups exhibited a similar inflammatory profile, which clustered between control and patient groups (Figure 6A). In serum, patient groups also segregated together and showed a very similar profile, which differed from the control group in the second principal component. The HCW groups clustered again between control and patient groups, with the Spn colonised HCWs appearing as a heterogeneous group (Figure 6B).

**Figure 6.**
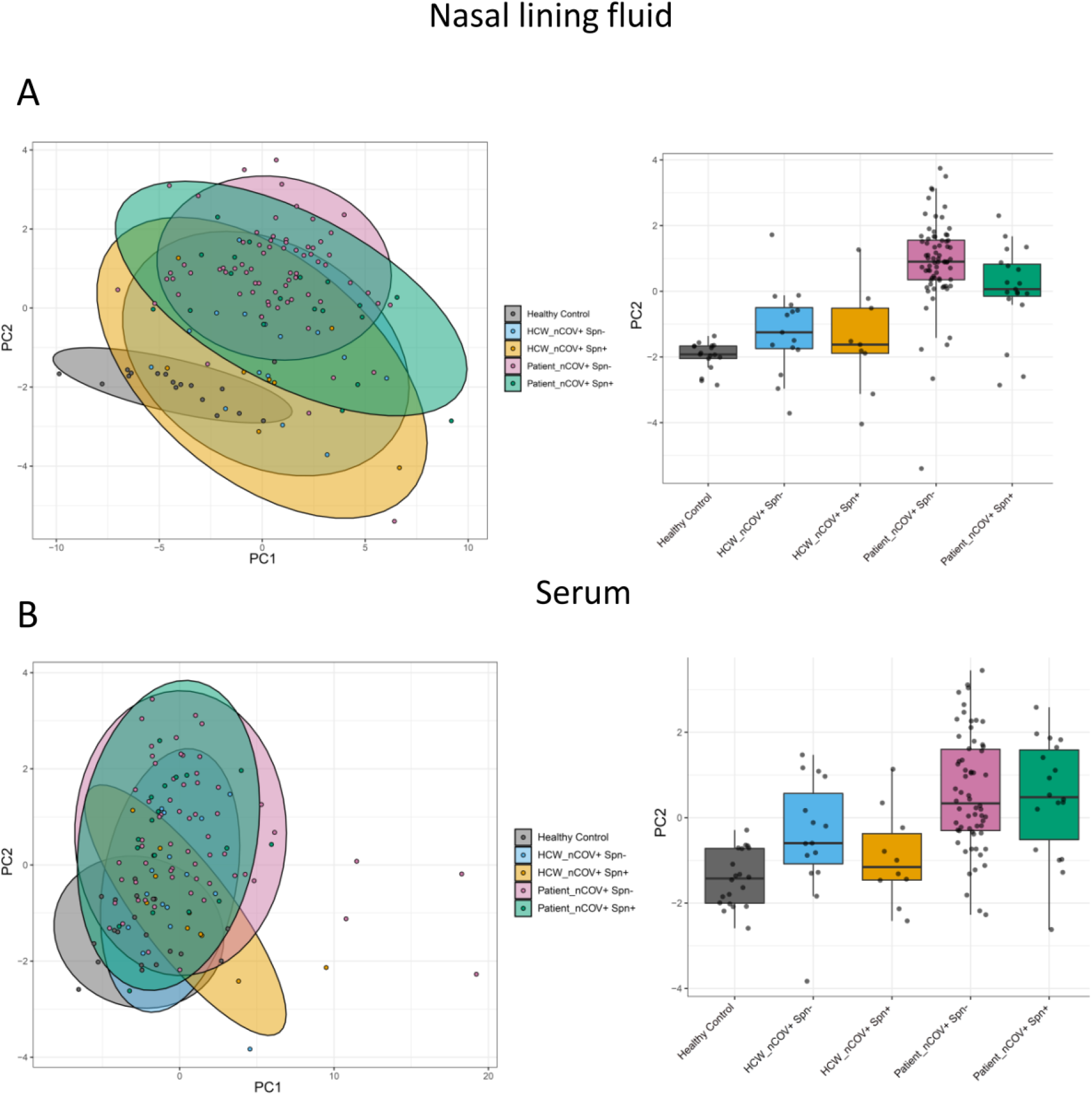
Nasal and serum inflammatory profiles of SARS-CoV-2 infection and co-infection with *S. pneumoniae* in HCWs and patients. A) Principal component analysis (PCA) of 30 cytokines in a) nasal lining fluid and b) serum of healthy control (grey), non-colonised HCW (light blue), Spn-colonised HCW (yellow), non-colonised patients (lilac) and Spn-colonised patient group (green). PC1: principal component 1; PC2, principal component 2.

## DISCUSSION

Here we report the first immunological analysis to our knowledge of SARS-CoV-2 infection in the context of co-infection with pneumococcus among two distinct cohorts - asymptomatic and mildly symptomatic HCWs and patients who experienced moderate to severe symptoms during SARS-CoV-2 infection. More importantly this is the first comprehensive analysis showing the potential role of *S. pneumoniae* carriage in modulating the host immune responses against SARS-CoV-2. Our findings have potential implications for other respiratory viruses.

We investigated mucosal and systemic antibody responses to the virus, memory B cell, CD4^+^ and CD8^+^ T cell anti-viral responses, as well as inflammatory responses and assessed whether colonising pneumococci modulate any of these arms of adaptive immunity or associate with an inflammatory profile early during the infection, making a number of important observations. Humoral and cellular antiviral immune responses varied substantially by pneumococcal carriage status both in HCWs who had asymptomatic to mild and patients who had moderate to severe SARS-CoV-2 infection, suggesting that colonisation of upper airways by pneumococcus may induce an immunosuppressive effect and thereby facilitating SARS-CoV-2 to escape protective host immune responses. This immunosuppressive effect was more apparent in the nasal mucosa of HCWs, where diminished salivary anti-S1 and S2 IgA levels were observed in the pneumococcal colonised individuals. In the patient cohort pneumococcal carriage was associated with reduced SARS-CoV-2 specific memory B cells and weakened T cell responses, particularly CD4^+^ T cell responses.

SARS-CoV-2 infection and virus replication starts in the naso/oropharynx-the primary site of infection^24^. Mucosal antibodies, particularly secretory IgA, play an important role in defence against respiratory viruses, as it promotes virus neutralisation and inhibits viral adhesion to the respiratory epithelium^25, 26^. For SARS-CoV-2 infection, *in vitro* studies with monoclonal anti-spike IgA demonstrated the superiority of IgA to block binding to the ACE2 receptor compared with the IgG isotype^27^. In addition, SARS-CoV-2 challenge studies in mice highlighted that musosal anti-spike IgA production is critical for sterilizing immunity in the upper respiratory tract^26^. We found that Spn colonised HCWs had significantly reduced IgA responses against SARS-CoV-2 S1 and S2 antigens compared to non-colonised individuals, and that these responses were similar in both Spn colonised and non-colonised groups in the patient cohort. This may indicate that such a suppressive effect is more important at the early phases of SARS-CoV-2 infection and less relevant once infection has progressed to symptomatic and severe disease.

We further investigated a potential mechanism responsible for the association between bacterial colonisation and IgA immunosuppressed responses against viral co-infections using the influenza virus infection model. *S. pneumoniae* and some other invasive respiratory pathogens (e.g., Neisseria species and *Haemophilus influenzae*) cleave human IgA1, which constitutes 90% of total IgA, by IgA1-proteases^17, 28^. This protease is primarily bacterial cell-associated but could potentially cleave also unbound, non-pneumococcal specific IgA1, contributing to the reduction in mucosal IgA to other pathogens, including SARS-CoV-2. Here, we expanded our previous observation to investigate the role of pneumococcal IgA1 protease in cleaving non-specifically anti-viral IgA. We observed that preceding experimental pneumococcal colonisation impaired the induction of both influenza-specific IgA1 and IgA2 nearly a month after influenza infection-an effect that was not seen in the absence of pneumococcal colonisation. This suggests that cleavage of IgA1 subclass by pneumococcal IgA1 protease most likely is not a mechanism, through which pneumococcus contributes to the reduction in mucosal IgA to other pathogens.

In addition, when influenza infection preceded experimentally induced pneumococcal colonisation, both Spn colonised and non-colonised, LAIV recipients mounted similar influenza-specific IgA levels. These findings imply that the order of exposure to respiratory pathogens during co-infection can affect some of the defence mechanisms. Differential nasal inflammatory responses during the early stages of infection, driven by either the virus or *S. pneumoniae,* depending on the order of infection, may have a differential effect on downstream immune responses^11, 15, 16^, altering the dynamics between the pathogens and fitness advantages^18^.

Recent reports demonstrated that SARS-CoV-2 virus is effective at avoiding or delaying the triggering of early innate immune responses, such as type I interferons (IFNs) *in vitro*^29^ and in humans^30, 31^. On the other hand, it has been shown that *S. pneumoniae* infection stimulates IFN-I production and upregulates the expression of IFN-stimulated genes in both mice and human studies^16, 32^. Therefore, it is possible that pneumococcal colonisation interferes with the replication cycle of the virus^33, 34^ and contributes to host antiviral defences by governing the production of IFNs^35, 36^. Here, despite a trend of higher viral load in the non-colonised groups, we did not observe significant viral load difference between pneumococcal colonised and non-colonised individuals. However, as SARS-CoV-2 viral load changes rapidly from day to day, the nature of the study prohibited the assessment of such a time-course dependent variable^37, 38^.

Consistent with published studies of COVID-19 infection^39, 40^, we observed inflammatory responses, including IL-6, IP-10, IL-1b and IL-8, in nasal lining fluid and serum in both cohorts, with increased cytokine induction in the patient groups, particularly the non-colonised individuals. The induction of cytokines that influence T cell activation (IL-2, IL-12, IFN-γ, IL-15, IL-17A), which subsequently assists B cell maturation ^41^, was distinctive in the nasal mucosa of patient cohort. Impairment of inflammatory response in the nasal mucosa could also affect the influx of effector immune cells and influence downstream immune responses ^11, 16^. Also, pneumococcal colonised individuals in both cohorts exhibited a lack of IL-2 and IL-12 induction, which could partially explain the weakened T cell responses observed in those groups.

The differences in the magnitude of immune responses between the two cohorts could be related to differences in viral load and severity of SARS-CoV-2 infection. Consistent with previous studies, convalescent patients mounted higher serum IgG levels compared to asymptomatic and mildly symptomatic HCWs ^42, 43^, showed increased frequency of memory B cells, broader and stronger T cell responses in the convalescent phase ^44^ and had elevated acute proinflammatory responses both in the nose and blood ^45^. Coordinated immunity by all three branches of adaptive immunity is more likely to protect against SARS-CoV-2 reinfection, as it is seen in protection against other infectious diseases^43^, whereas suboptimal immunity against SARS-CoV-2 could allow reinfection to occur.

The differential prevalence of pneumococcal carriage between COVID-19 cases and unexposed individuals amongst HCWs supports the hypothesis of pneumococcus and SARS-CoV-2 interplay in the upper respiratory tract. Co-infections amongst COVID-19 cases in HCWs likely reflect bacterial and viral transmission prior to social distancing and other community interventions. These observations should be evaluated in larger epidemiological studies, providing also the opportunity to assess whether the pneumococcus and SARS-CoV-2 interrelationship is serotype-specific, as has been previously reported in mouse and epidemiological studies of other viral co-infections, such as influenza and RSV^46–48^.

Our study has limitations. Diagnosis of co-infection was complex amongst patients, as pneumococcus might be carried by the patient before the viral infection or might be picked up later. High use of antibiotics (potentially prescribed at an outpatient visit) and reduced social mixing affected survival, prevalence and transmission dynamics of other respiratory pathogens, such RSV^1, 49^, and most likely pneumococcal transmission. Hence, we observed decreased prevalence of pneumococcus amongst patients-particularly during periods of national lockdown, which subsequently limited the number of SARS-CoV-2/ pneumococcus co-infected individuals studied here. Thus, despite inclusion of nearly 500 subjects in our study, we were able to evaluate pneumococcal mediated immunomodulatory effects only for a relatively small number of individuals, limiting the ability to do further stratifications.

In children, pneumococcal colonisation has been profusely documented. Children however experience an attenuated course of SARS-CoV-2 infection. This emphasises the importance of our findings in adults and indicate the need to conduct future studies aimed at understanding the differential disease course in children, taking this important variable into consideration. In addition, differences in age and underlying disease between the HCW and patient cohorts are some factors that have potentially affected the course and outcome of the disease. Further studies, ideally in the setting of controlled human co-infection model, are needed to explore S*. pneumoniae*/respiratory virus interactions and the biological mechanisms through which pneumococcus assists viruses to subvert immune responses at the primary site of infection.

Despite the observational design, our study has identified pneumococcal colonisation as an important variable which can modulate host immune responses to SARS-CoV-2 infection; an effect that was observed in both cohorts despite the aforementioned differences. An impaired adaptive immunity, including all three arms of immune responses, against SARS-CoV-2 natural infection could potentially increase susceptibility to subsequent SARS-CoV-2 infection or associate with long-Covid symptoms. The increased evidence on PCV-induced protection against lower respiratory infections associated with viral infection and the broader ability of pneumococcus to interact with respiratory viruses in a way that increases pneumococcal virulence, viral pathogenicity or impairs anti-viral immune responses highlights the importance of PCVs in both paediatric and older adults as an additional public health tool for those who are at increased risk of pneumococcal and viral lower respiratory infections.

## METHODS

### Ethical approval and participant recruitment

This study combined participants recruited into two prospective cohort studies of a) frontline HCWs (n=85) and b) patients (n=400) presented to the hospital. HCWs in a variety of roles were enrolled onto SARS-CoV-2 Acquisition in Frontline Healthcare Workers - Evaluation to inform Response (SAFER) study between 30^th^ March and 9^th^ April 2020 at Royal Liverpool University Hospital (RLUH) in Liverpool, UK. Eligible HCWs were over the age of 18 years, asymptomatic at the time of enrolment to the study and were working in a patient facing role for at least one day during the study period (12 weeks from enrolment). Screening against SARS-CoV-2 and *S. pneumoniae* was performed on the nasopharyngeal and saliva samples (Figure 1SA) and symptom reporting was via a questionnaire completed twice weekly, accompanying each sampling episode^19^.

For the patient cohort, adults (aged ≥ 18 years) with signs and symptoms of suspected COVID-19 infection attending RLUH, Aintree University and Whiston Hospital in Merseyside between April 2020 to January 2021 were recruited into Facilitating A SARS Cov-2 Test for Rapid Triage (FASTER), regardless of disease severity, race, ethnicity, gender, pregnancy or nursing status, or the presence of other medical conditions. Study exclusion criteria included lack of willingness or ability to provide informed consent, or lack of an appropriate legal guardian or representative to provide informed consent.

The two study protocols were reviewed and approved by the NHS Health Service Research Ethics Committees (REF: 20/SC/0147 for SAFER and 20/SC/0169 for FASTER). All participants provided written informed consent and were free to withdraw from the studies at any point.

### Immunologic Analyses of SARS-CoV-2 positive Participants

Individuals from both cohorts (all Spn colonised individuals and a subset of non-colonised patients) who were SARS-CoV-2 positive were stratified by pneumococcal colonisation status, and pre-pandemic samples from healthy unexposed individuals were also included, resulting in 5 groups used for the immunological analysis: i) HCW_nCOV+ Spn-, ii) HCW_nCOV+/Spn+, iii) Patient_nCOV+/Spn-, iv) Patient_nCOV+/Spn+ and v) Healthy controls. To assess and compare immune responses to SARS-CoV-2, we analysed convalescent blood and upper respiratory samples (saliva and nasal lining fluid) in HCW and patient groups. Access to convalescent samples was restricted in the patient cohorts, as only 39% (100/254) of SARS-CoV-2 positive individuals complied with the study protocol and donated samples at the convalescent phase of COVID-19 infection. In both cohorts, we also assessed nasal and systemic inflammation during the acute phase of COVID-19 infection. Samples from healthy adults collected prior to June 2019 were used as healthy controls. Demographic and clinical characteristics of these 5 groups are shown in Table 1.

### Sample collection

HCWs were asked to provide a self-collected flocked combined nasopharyngeal swab (Amies, MWE, UK) twice per week, in addition to two saliva samples (raw material and in 1ml of STGG) into sterile tubes (STARSTED, USA) and two SAM filters (Nasosorption FX, UK) samples, collected once per week. Peripheral blood samples were collected by the Clinical Research Unit team. Serum samples were collected on the recruitment day and monthly during the 12weeks follow-up, whereas PBMCs were collected on the day of recruitment and at 3-months visit.

For the patient cohort, recruitment was either in the A&E unit or in wards within 48h of hospital admission. Whole blood, respiratory samples (nose and throat swabs, oral swabs, saliva in STGG, SAM filters) and urine were collected at the point of patient recruitment. If the patient had a positive SARS-CoV-2 test and stayed in the hospital, then day-2 and a day-7 samples (from study recruitment day) were collected. All patient participants were invited to visit the Accelerator Research Clinic at Liverpool School of Tropical Medicine and donate samples at the convalescent phase (Day28 ± 25 days from recruitment day). Convalescent patients had to be symptom-free and approximately 3 weeks out from symptoms onset.

For healthy controls, we used samples collected from healthy adults (18-73 years of age) who participated into one of our experimental human pneumococcal carriage studies. These were baseline (prior to inoculation with pneumococcus) blood and respiratory samples, collected between 2015 to 2019 and were considered to be unexposed controls given that SARS-CoV-2 emerged as a novel pathogen in late 2019 (November to December).

### PBMC isolation and handling

Whole blood was collected in lithium heparin tubes (BD vacutainer, USA) for all study participants (HCWs, patients and healthy, unexposed donors) and PBMCs were isolated using density-gradient sedimentation in a biosafety level 3 facility. To isolate PBMCs, blood 1:1 diluted in PBS was layered over Ficoll-Paque in a SepMate tube (HCWs and patients) or Leucosep tubes with porous barriers (healthy donors) and centrifuged for 10 min at 1,200*g*. The PBMC layer was collected by quickly pouring the content into a new 50-ml tube. Isolated PBMCs were then cryopreserved in CTL-Cryo ABC media kit (Immunospot, Germany) and frozen overnight at -80°C, and then transferred to liquid nitrogen until further analysis.

### Serum isolation and handling

Whole blood was collected in serum separator tubes (SST BD Vacutainer tubes, USA) and centrifuged for 10min at 1200g for serum separation in a biosafety level 3 facility. The serum was then aliquoted and stored at -80°C until further use. Prior to use in assays, serum samples were treated with 1% Triton-X-100 for 1h in room temperature^50^.

### Respiratory samples handling

NP swabs collected from patients were transferred to LSTM diagnostic laboratory and processed immediately. Self-collected NP swabs from HCWs were stored at -80°C till further use. Throat swabs in STGG, saliva raw and saliva in STGG, SAM filters were stored at -80°C till further use. Where appropriate, respiratory secretion samples were treated with 1% Triton-X-100 for 1h at room temperature prior to use in SARS-CoV-2 ELISAs or cytokine beads assay.

### Bacterial DNA extraction and S. pneumoniae qPCR

Bacterial genomic DNA was extracted from raw and culture-enriched (CE) samples, as previously described ^51^. Briefly, in the patient cohort, DNA was extracted from 350μl raw nose to throat swabs in Amies plus 50% STGG (skim milk, tryptone, glucose, glycerine), 350μl raw oropharyngeal swabs (OPS) in 50% STGG and 200μL raw saliva samples. In the HCW study, DNA was extracted from 350μl raw nose to throat swabs in Amies and 200μL raw saliva samples. Raw aliquots were stored at -20°C until the next day. For culture-enrichment, 50μl of raw sample was cultured on Columbia blood agar supplemented with 5% horse blood (PB0122A, OXOID/Thermo Scientific) and 80μl gentamycin 1mg/mL (G1264-250mg, Sigma-Aldrich co Ltd). Plates were incubated overnight at 37°C and 5% CO_2_. The remaining sample was stored at -80°C. The next day, all bacterial growth was harvested into 2mL 50% STGG and vigorously vortexed until homogenised. Bacterial DNA from both raw and culture enriched material was extracted using the Agowa Mag mini-DNA extraction kit (LGC Genomics, Berlin, Germany).

### Spn qPCR (gene targets, thresholds, positive controls etc)

Pneumococcal presence was determined by sequential singleplex qPCR targeting the *lyt*A ^52^ and the *pia*B genes ^53^, using the QuantStudio 5 system (ThermoFisher, UK), as previously described ^53^. Briefly, 20μL PCR mix consisted of 12.5μL 1 × TaqMan Universal PCR Master Mix (Life Technologies Ltd, Paisley, UK), 0.225μL or 0.2μL *100μ*M each *lytA* or *piaB* primer respectively, 0.125μLor 0.175μL 100μM *lytA* or *piaB* probe respectively, molecular graded water (Fisher Scientific, Loughborough, UK) and 2.5μL of the extracted DNA. Thermal cycling conditions were: 10min at 95°C and 40 cycles of 15secs at 95°C and 1min at 60°C. A negative DNA extraction control (parallel extraction from sample buffer only), a qPCR negative control (master mix only), a qPCR positive control (pneumococcal Spn15B strain) and duplicates of each sample were amplified. A standard curve of a ten-fold dilution series of genomic DNA extracted from TIGR4 was used. Pneumococcal positive samples considered those that both genes were present. All samples were assessed by a *lytA* qPCR and those positive underwent a *piaB* qPCR. Samples were considered *lytA*-positive if one or two yielded a C_T_ < 40 cycles. Threshold between plates was normalised according to positive control C_T_ values.

### SARS-CoV-2 RNA extraction and RT-qPCR

SARS-CoV-2 RNA was extracted from 140 µl amies solution using the Qiagen Viral RNA Mini Kit (Qiagen, Germany) following manufacturer’s instructions. In addition, the internal extraction control from the qPCR kit was added to each sample prior to extraction as per manufacturer’s instructions. For SARS-CoV-2 RT-qPCR detection, 8μl of extracted RNA was tested using the Genesig® Real-Time Coronavirus COVID-19 PCR assay (Genesig, UK) in a RGQ 6000 thermocycler (Qiagen, Germany). Samples were classified as RT-qPCR positive if both the internal extraction and the SARS-CoV-2 probes were detected at <40Ct. Virus copies/ml were quantified using the manufacturer’s positive control (1.67 x 105 copies/μl) as a reference.

### Enzyme-linked immunosorbent assay for SARS-CoV-2 and influenza virus antigens

ELISA was used to quantify levels of IgG and IgA to SARS-CoV-2 antigens in serum and saliva or nasal lining fluid samples, respectively, whereas IgA, IgA1 and IgA2 were measured in nasal wash samples of LAIV recipients. Recombinant SARS-CoV-2 receptor binding domain (RBD) and nucleocapsid protein were produced at the University of Oxford, as reported elsewhere ^54^. Recombinant S1 and S2 subunits (full length proteins) were commercially available (Biosciences, UK). Seasonal TIV (either 2015/2016 or 2016/2017 formulation) was used as the source of influenza antigens for measuring mucosal IgA and subclasses in nasal washes of study participants.

Antibody levels to SARS-CoV-2 and influenza antigens were quantified, as previously described ^16, 55^ with minor modifications. Briefly, Nunc 96-well plates were coated with 1μg/ml SARS-CoV-2 antigen or 0.2 μg/ml TIV and stored at 4°C overnight for at least 16h. After coating, plates were washed 3 times with PBS/0.05%Tween and blocked with 2% BSA in PBS for 1h at room temperature. Thawed serum, saliva, nasal fluid and nasal wash samples diluted in 0.1% BSA-PBS were plated in duplicate and incubated for 2h at room temperature alongside an internal positive control (dilution of a convalescent serum) to measure plate to plate variation. For the standard curve, a pooled sera of SARS-CoV-2 infected participants was used in a two-fold serial dilution to produce either eight or nine standard points (depending on the antigen) that were assigned as arbitrary units. Goat anti-human IgG (γ-chain specific, A9544, Sigma-Aldrich) or IgA (α-chain specific, A9669, Sigma-Aldrich) or mouse anti-human IgA1 (Fc-specific, ab99794, Abcam) or IgA2 (Fc-specific, ab99800, Abcam) conjugated to alkaline phosphatase was used as secondary antibody and plates were developed by adding 4-nitrophenyl phosphate in diethanolamine substrate buffer. Optical densities were measured using an Omega microplate reader at 405nm. Blank corrected samples and standard values were plotted using the 4-Parameter logistic model (Gen5 v3.09, BioTek).

### Flow Cytometry

#### PBMCs preparation for flow cytometry and stimulation assays

Cryopreserved cells were thawed by diluting them in 10ml pre-warmed RPMI, containing 10% FBS and 1% PNS in presence of benzonase and spun at 200g for 10 min. Supernatant was carefully removed and the process was repeated once again. Pelleted cells were resuspended in warm medium, counted and apportioned for the assays.

#### Direct *ex vivo* immune B and T cell phenotyping

<u>B cell phenotyping</u>: Cryopreserved human PBMC in 96-well U plates were washed (440g for 5 min), stained with Live/dead e506 viability dye for 15 minutes at 4°C protected from light (Thermofisher), and an extracellular cocktail of monoclonal antibodies, including SARS-CoV-2 Spike-1 and -2 protein conjugated with Biotin-Streptavidin, for 20 minutes at 4°C protected from light (Table S1). SARS-CoV-2 S1 and S2 proteins were conjugated with Biotin (EZ Link conjugation kit, Thermofisher), then directly labelled with Streptavidin-BV785 and PE (Biolegend), respectively. Cells were washed again and resuspended in 200µL of PBS, in each well. All samples were acquired on an Aurora cytometer (Cytek Biosciences) and analysed using Flowjo software v.10 (Treestar).

<u>T cell phenotyping:</u> Following stimulation in 96-well U plates. PBMC cells were washed (440g for 5 min), stained with Live/dead e506 viability dye for 15min at 4°C protected from light (Thermofisher), and an extracellular cocktail of monoclonal antibodies for 20 minutes at 4°C protected from light (Table S2). Cells were washed again (440g for 5 min), then fixed and permeabilized with CytoFix/CytoPerm (BD Biosciences) for 15min at 4oC. After the incubation, cells were stained with an intracellular cocktail of monoclonal antibodies (Table S2). Following incubation, cells were washed and resuspended in 200µL of PBS per well. All samples were acquired on an Aurora cytometer (Cytek Biosciences) and analysed using Flowjo software v.10 (Treestar).

### T cell stimulation and Intracellular cytokine staining assay

Cells were cultured for 18h at 37°C in the presence of SARS-CoV-2 specific peptides (2μg/ml) in 96-well U bottom plates at 1×10^6^ PBMCs per well in complete media. Overlapping peptides spanning the immunogenic domains of the SARS-CoV-2 Spike (Prot_S), nucleocapsid (Prot_N) and S1 subunit (Prot_S1) proteins were purchased from Miltenyi Biotec. Golgi-Plug containing brefeldin A and golgi-stop containing monensin (BD Biosciences, San Diego, CA) were added 2h after the peptide addition. A stimulation with an equimolar amount of DMSO was performed as a negative control and Staphylococcal enterotoxin B (SEB, 2 μg/mL) was included as a positive control. The following day cells were harvested from plates, washed and stained for surface markers (Table S2).

### Luminex analysis of nasal lining fluid or serum

The Cytokine Human Magnetic 30-plex panel was used to quantitate human nasal and serum cytokines. The panel included the following 30 analytes: G-CSF, GM-CSF, IFN-α, IFN-γ, IL-1β, IL-1RA, IL-2, IL-2R, IL-4, IL-5, IL-6, IL-7, IL-8, IL-10, IL-12 (p40/p70), IL-13, IL-15, IL-17A, TNF-α, Eotaxin, IP-10, MCP-1, MIG, MIP-1α, MIP-1β, Rantes, EGF, FGF-b, HGF and VEGF). Cytokines were eluted from stored SAM filters using 100 μl of assay diluent buffer (Thermo Fisher Scientific) by centrifugation, then the eluate was cleared by further centrifugation at 16,000*g*, following an additional centrifugation at 16,000*g* for 10 min to clear samples. Triton-treated nasal fluid and serum were acquired on an LX200 using a 30-plex magnetic human Luminex cytokine kit (Thermo Fisher Scientific) and analysed with xPonent3.1 software following the manufacturer’s instructions. Samples were run in duplicate, and standards run on all plates. Calibration and verification beads were run prior to all runs.

### Statistical Analysis

Statistical analyses were performed using R software (version 4.0.4) or GraphPad Prism (version 9.0). Two-tailed statistical tests were used throughout the study. Clinical and demographic data were analysed using GraphPad. Categorical variables were compared using Fisher’s exact or Chi-squared test. Continuous variables were tested for normality and appropriate statistical tests applied. Non-normally distributed measurements were expressed as the median and Mann-Whitney (two group comparison) or Kruskal-Wallis (three to five group comparison) tests were used. Differences were considered significant at p < 0.05, and multiple testing correction was employed where appropriate. For volcano plots and correlograms, false discover rate (FDR) corrections were performed using the Benjamini-Hochberg test at an FDR < 0.05 significance threshold. Correlations were assessed using Pearson’s correlation test using either raw or log-transformed values.

## Data Availability

LSTM repository

## Acknowledgements

We acknowledge and thank all the participants recruited to the SAFER and FASTER clinical studies. We also thank the LUHFT Clinical Research Unit and NIHR research nurses, who assisted with sample collection. We thank Ben Dugan, Dessi Loukov, Chris Myerscough, Natalie Tate, Alexander Tinworth and the LSTM Diagnostic team for the excellent technical and logistical support. We also thank Dr Florian Krammer (Icahn School of Medicine at Mount Sinai) for providing RBD plasmids for protein production. We acknowledge and thank University College London Hospitals NHS Foundation Trust for sponsorship of the SAFER study. S.J.D. is a Jenner Investigator and held a Wellcome Trust Senior Fellowship [106917/Z/15/Z].

## Author contributions

EM: conception and design of the study, protocol development, assay development, data analysis and interpretation and manuscript writing. JR and BCU: assay development, conduction of experiments and data analysis. CS, EN, SP, AH, LH, ELG, KSC, RLB, CTW, ACA contributed to sample processing, conducting, and analysing experiments. AHW, SG, MF, KL, HL, AMC, NFW, TF and ERA contributed to recruitments, samples collection and design of the study. ACA, SJD, DP reviewed and commented on the manuscript. RB: study oversight, protocol development, review and comment on the manuscript. EB: study oversight, protocol development, data interpretation, review and comment on the manuscript. CT: data interpretation, review and comment on the manuscript. LJ, BDG and DMF: conception and design of the study, protocol development, data interpretation and manuscript writing. All authors have read and approved the manuscript.

