## Supplemental material for "*Streptococcus pneumoniae* colonisation associates with impaired adaptive immune responses against SARS-CoV-2"

**Supplementary material**

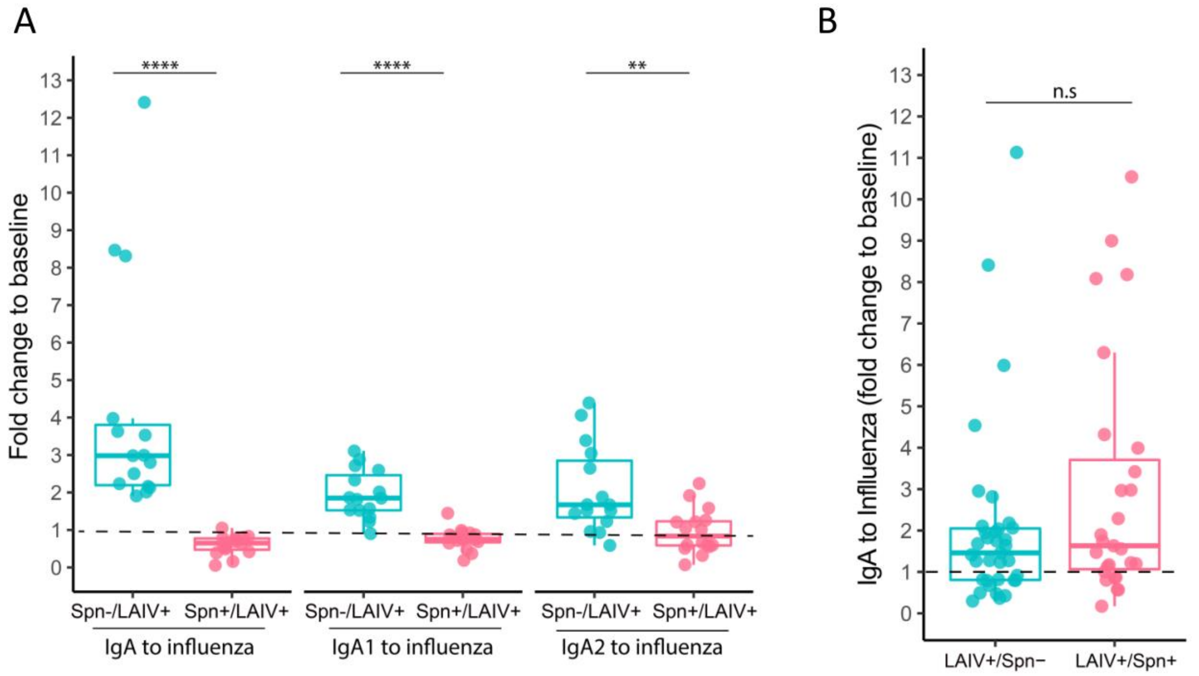


**Figure S1: Mucosal IgA antibodies against influenza antigens**. A) Fold change (day 24 post LAIV administration versus baseline) in nasal levels of IgA, IgA1 and IgA2 to influenza antigen stratified by pneumococcal status (Spn-/LAIV+ in light blue, n=15 and Spn+/LAIV+ in light red, n= 15) in individuals challenged with live pneumococcus 3 days before LAIV administration. B) Fold change (day 27 post LAIV administration versus baseline) of nasal IgA to influenza antigens in individuals challenged with live pneumococcus 3 days after LAIV administration (LAIV+/Spn-, n=35 and LAIV+/Spn+, n=27). Medians with IQRs are depicted and each spot represents an individual. **p<0.01, ****p < 0.0001 by Mann-Whitney test comparing fold-change levels between carriage– and carriage+ subjects.

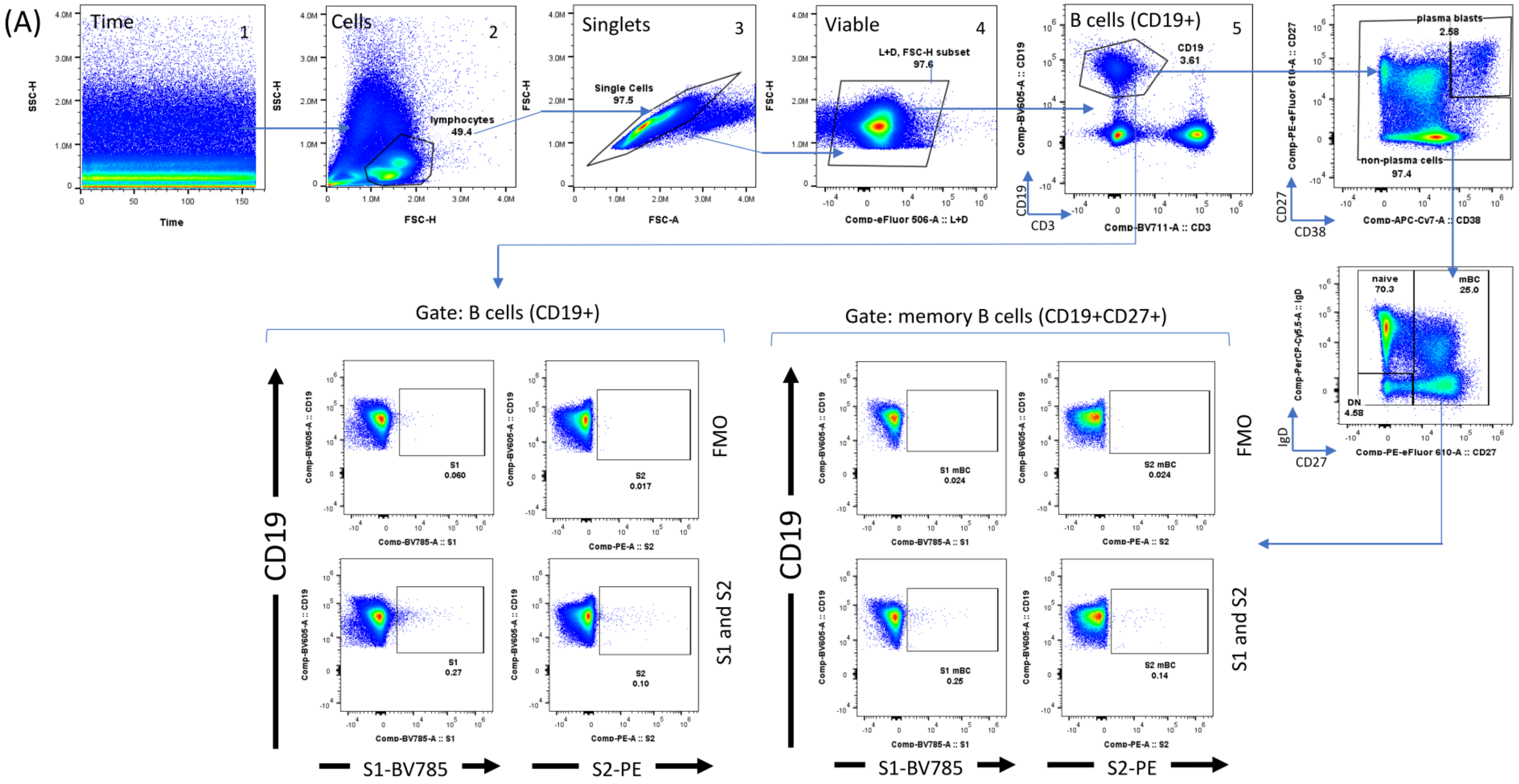


**Figure S2. Gating strategy for B cell subset analysis using a representative PBMC sample.** Identification of the B cells based on the following steps from left to right: 1) Time dot-plot to determine the quality of stability of the acquisition 2) Adequate adjustment of the gate of lymphocytes 3) Exclusion of doublets with the identification of singlets improving the accuracy of the analysis 4) Selection of the viable lymphocytes 5) Identification of the B cells based on CD19+ expression. Analysis within B cells (CD19+) subset. Subsequently, we identified plasma blasts (CD27+CD38++) and non-plasma cells (CD38-) based on the expression of CD27 (memory cell marker) vs. CD38 (plasma cell marker). From non-plasma cells we identified the naïve, memory B cells (mBC) and the double negative population based on the expression of IgD vs. CD27. Finally, to delineate the SARS-CoV-2 specific B cells against S1 and S2, we analysed within the B cells (CD19+) and memory B cells (mBC) the percentage of expression S1 and S2 proteins conjugated with biotin and labelled with Streptavidin (BV785 and PE, respectively). Abbreviations: FMO (Fluorescence Minus One).

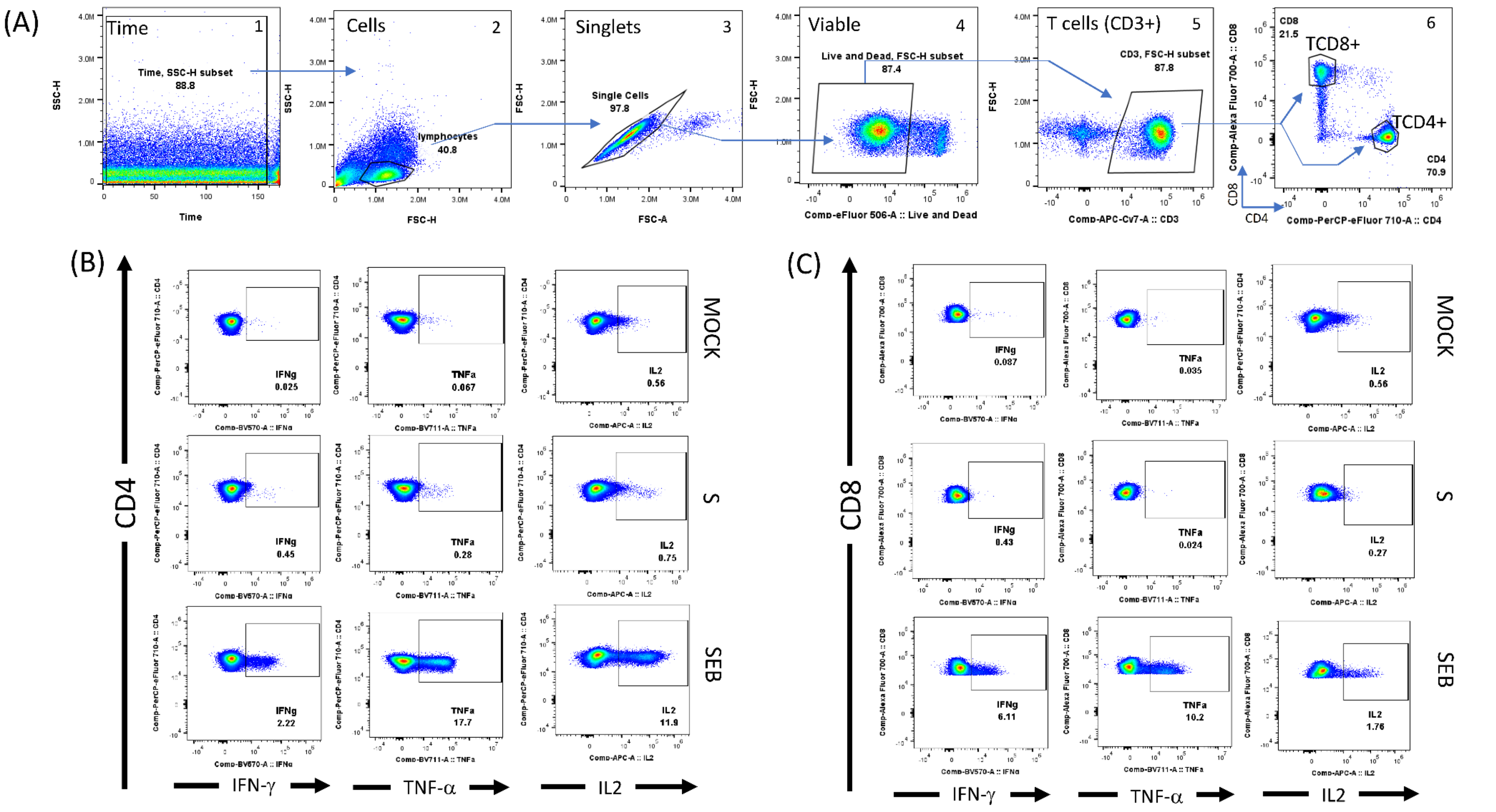


**Figure S3. Gating strategy for T cell subset analysis (CD4+ and CD8+) by flow cytometry using a representative PBMC sample. A)** Identification of the T cellular subsets CD4 and CD8 based on the following steps: 1) Time dot -plot to determine the quality of stability of the acquisition 2) Adequate adjustment of the gate of lymphocytes 3) Exclusion of doublets with the identification of singlets improving the accuracy of the analysis 4) Selection of the viable lymphocytes 5) Identification of the T cells based on CD3+ expression 6) Identification of the TCD4+ and TCD8+ within CD3+ cells. **B)** Analysis within TCD4+ cells subset. Representative dot-pots of cytokine production (IFN-$\gamma$, TNF-$\alpha$ and IL2) following SARS-CoV-2 Spike (S) protein stimulation (2µg/mL) and SEB (Staphylococcus Enterotoxin B) as positive control compared to mock (unstimulated) within CD4+ cells. TCD4+ cells were also stimulated SARS-CoV-2 with S1 subunit (S1) and Nucleocapsid (N) (not shown) (C) Analysis within TCD8+ cells subset. Representative dot-pots of cytokine production (IFN-$\gamma$, TNF-$\alpha$ and IL2) following SARS-CoV-2 Spike protein stimulation (2µg/mL) and SEB (Staphylococcus Enterotoxin B) as positive control compared to mock (unstimulated) within CD8+ cells. CD8+ cells were also stimulated SARS-CoV-2 with S1 subunit (S1) and Nucleocapsid (N) (not shown).

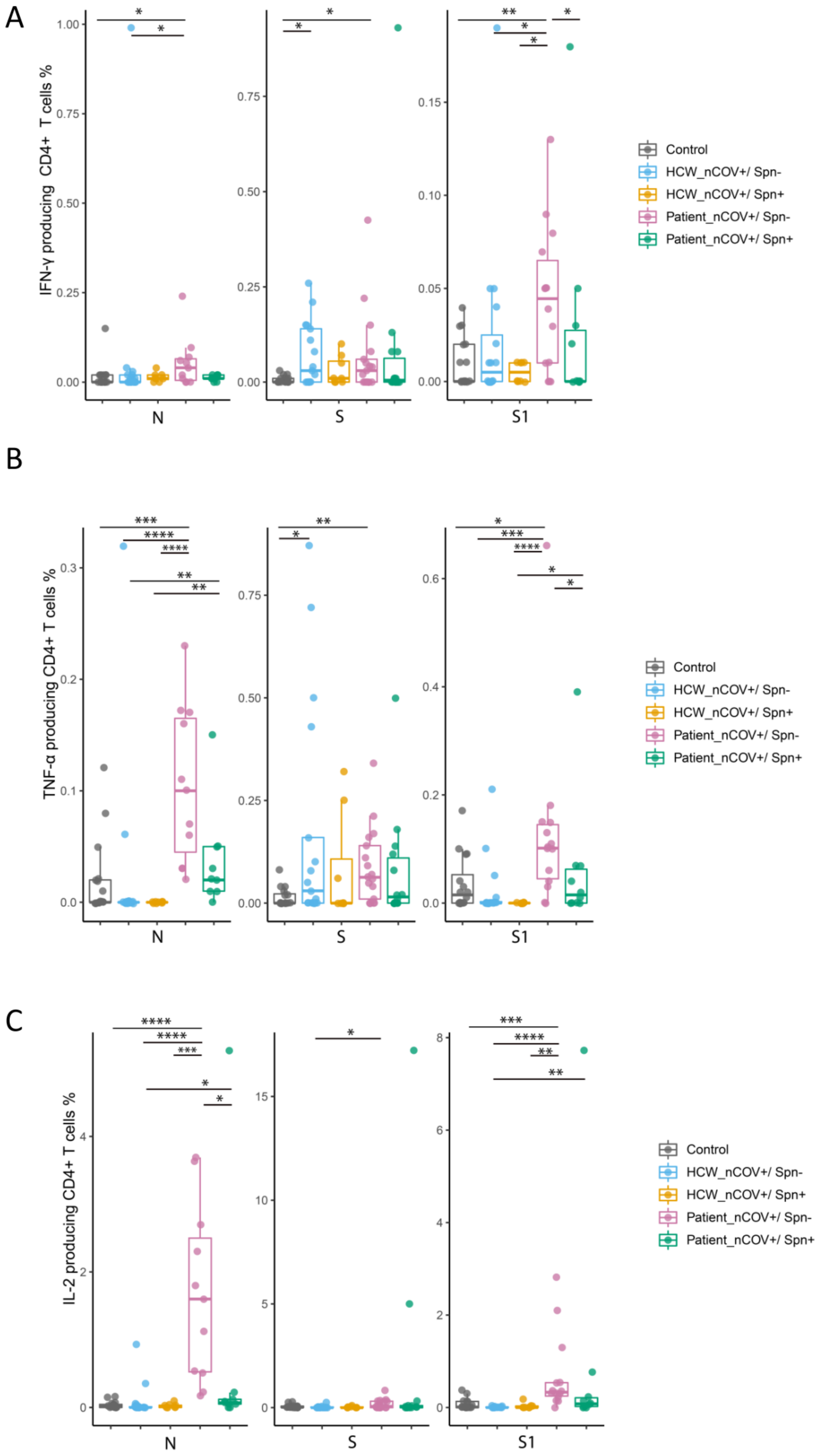

A
IFN-γ producing CD4+ T cells %
N
S
S1
Control
HCW_nCOV+/ Spn-
HCW_nCOV+/ Spn+
Patient_nCOV+/ Spn-
Patient_nCOV+/ Spn+
B
TNF-α producing CD4+ T cells %
N
S
S1
Control
HCW_nCOV+/ Spn-
HCW_nCOV+/ Spn+
Patient_nCOV+/ Spn-
Patient_nCOV+/ Spn+
C
IL-2 producing CD4+ T cells %
N
S
S1
Control
HCW_nCOV+/ Spn-
HCW_nCOV+/ Spn+
Patient_nCOV+/ Spn-
Patient_nCOV+/ Spn+

**Figure S4.** Percentage of A) IFN-γ, B) TNF-α and C) IL-2 producing CD4+ T cells after ex vivo PBMC stimulation with N, S1 and S peptides pools in exposed HCWs (non-colonised; n=17 and Spn-colonised; n=8), recovered patients (non-colonised; n=17 and Spn-colonised; n=14) and healthy control (n=16). One peptide pool was used per condition. SEB was used as a positive control and DMSO as the negative control (non-stimulated cell condition-mock). Background (mock) was subtracted from peptide-stimulated conditions to remove non-specific signal. Medians with IQRs are depicted and each spot represents an individual. *p <0.05, **p<0.01, ***p < 0.001, ****p < 0.0001 by Kruskal-Wallis test.

| Marker | Fluorochrome | Clone | Isotype | Provider | Reference |
|---|---|---|---|---|---|
| CD19 | BV605 | HIB19 | Mouse IgG1, κ | Biolegend | 302244 |
| CD3 | BV711 | SK7 | Mouse IgG1, κ | Biolegend | 344838 |
| CD27 | PE-eF610 | O323 | Mouse IgG1, κ | Thermofisher | 61-0279-42 |
| IgD | PerCP-Cy5.5 | IA6-2 | Mouse IgG2a, κ | Biolegend | 348208 |
| CD38 | APC-Cy7 | HIT2 | Mouse IgG1, κ | Biolegend | 303534 |
| S1 | Strep-BV785 | NA | NA | Biolegend | 405249 |
| S2 | Strep-PE | NA | NA | Biolegend | 405203 |
| Live & Dead | e506 | NA | NA | Thermofisher | 65-0866-14 |

**Supplemental table 1. Summary and specifications of the multiparametric flow cytometry panel used for the B cell analysis by flow cytometry**. We developed a multiparametric flow cytometry panel composed with different monoclonal antibodies (from Biolegend and BD Biosciences) including a viability dye (Thermofisher), and SARS-CoV-2 S1 and S2 proteins conjugated with biotin (EZ Link, Thermofisher) and labelled with Streptavidin BV785 and PE (Biolegend), respectively. Electronic compensation was set using CompBeads (BD Biosciences), Arc beads (ThermoFisher) and using the Spectral Flow automated unmixing software (Aurora, Cytek Biosciences) according to manufacturer's instructions. Abbreviations: NA, Not applicable.

| Marker | Fluorochrome | Clone | Isotype | Provider | Reference |
|---|---|---|---|---|---|
| CD3 | APC-Cy7 | SK7 | Mouse IgG1, κ | Biolegend | 344818 |
| CD4 | PerCP-e710 | SK3 | Mouse IgG1, κ | Thermofisher | 46-0047-42 |
| CD8 | AF700 | SK1 | Mouse IgG1, κ | Biolegend | 344724 |
| Live & Dead | e506 | NA | NA | Thermo | 65-0866-14 |
| IFN-γ | BV570 | 4S.B3 | Mouse IgG1, κ | Biolegend | 502534 |
| TNF-α | BV711 | MAb11 | Mouse IgG1, κ | Biolegend | 502940 |
| IL-2 | APC | MQ1-17H12 | Rat IgG2a, κ | Biolegend | 500307 |

**Supplemental table 2. Summary and specifications of the multiparametric flow cytometry panel used for T cell analysis by flow cytometry**. We developed a multiparametric flow cytometry panel composed with extracellular (CD3, CD4 and CD8) and intracellular (cytokines) monoclonal antibodies (from Biolegend and BD Biosciences) including a viability dye (Thermofisher) to assess SARS-CoV-2 specific T cells in HCW and patients with SARS-CoV-2 infection by flow cytometry. Electronic compensation was set using CompBeads (BD Biosciences), Arc beads (ThermoFisher) and using the Spectral Flow automated unmixing software (Aurora, Cytek Biosciences) according to manufacturer's instructions. Abbreviations (NA, Not applicable).
